# Coagulation factors and COVID-19 severity: Mendelian randomization analyses and supporting evidence

**DOI:** 10.1101/2020.11.20.20235440

**Authors:** Yao Zhou, Zipeng Liu, Hongxi Yang, Jianhua Wang, Tong Liu, Kexin Chen, Yaogang Wang, Pak Chung Sham, Ying Yu, Mulin Jun Li

## Abstract

**Background:** The evolving pandemic of COVID-19 is arousing alarm to public health. According to epidemiological and observational studies, coagulopathy was frequently seen in severe COVID-19 patients, yet the causality from specific coagulation factors to COVID-19 severity and the underlying mechanism remain elusive.

**Methods:** First, we leveraged Mendelian randomization (MR) analyses to assess causal relationship between 12 coagulation factors and severe COVID-19 illness based on two genome-wide association study (GWAS) results of COVID-19 severity. Second, we curated clinical evidence supporting causal associations between COVID-19 severity and particular coagulation factors which showed significant results in MR analyses. Third, we validated our results in an independent cohort from UK Biobank (UKBB) using polygenic risk score (PRS) analysis and logistic regression model. For all MR analyses, GWAS summary-level data were used to ascertain genetic effects on exposures against disease risk.

**Results:** We revealed that genetic predisposition to the antigen levels of von Willebrand factor (VWF) and the activity levels of its cleaving protease ADAMTS13 were causally associated with COVID-19 severity, wherein elevated VWF antigen level (*P* = 0.005, odds ratio (OR) = 1.35, 95% confidence interval (CI): 1.09-1.68 in the Severe COVID-19 GWAS Group cohort; *P* = 0.039, OR = 1.21, 95% CI: 1.01-1.46 in the COVID-19 Host Genetics Initiative cohort) and lowered ADAMTS13 activity (*P* = 0.025, OR = 0.69, 95% CI: 0.50-0.96 in the Severe COVID-19 GWAS Group cohort) lead to increased risk of severe COVID-19 illness. No significant causal association of tPA, PAI-1, D-dimer, FVII, PT, FVIII, FXI, aPTT, FX or ETP with COVID-19 severity was observed. In addition, as an independent factor, VWF PRS explains a 31% higher risk of severe COVID-19 illness in the UKBB cohort (*P* = 0.047, OR per SD increase = 1.31, 95% CI: 1.00-1.71). In combination with age, sex, BMI and several pre-existing disease statues, our model can predict severity risks with an AUC of 0.70.

**Conclusion:** Together with the supporting evidence of recent retrospective cohort studies and independent validation based on UKBB data, our results suggest that the associations between coagulation factors VWF/ADAMTS13 and COVID-19 severity are essentially causal, which illuminates one of possible mechanisms underlying COVID-19 severity. This study also highlights the importance of dynamically monitoring the plasma levels of VWF/ADAMTS13 after SARS-CoV-2 infection, and facilitates the development of treatment strategy for controlling COVID-19 severity and associated thrombotic complication.

## Introduction

The outbreak of coronavirus disease 19 (COVID-19), which is caused by severe acute respiratory syndrome coronavirus 2 (SARS-CoV-2), with an unprecedented number of pneumonia cases from the late December 2019 put people on full alert ^1^. Widespread comorbidities implicating several organs were frequently observed in COVID-19 patients, such as diseases in cardiovascular, neurological and hematopoietic systems ^2^. Of note, COVID-19-associated coagulopathy is a common complication among those patients developing severe systemic diseases and multiorgan failure ^3-5^, suggesting the importance of exploring clinical markers and the causal association between coagulopathy and COVID-19 ^6^.

Some coagulation parameters including D-dimer, prothrombin time (PT), von Willebrand factor (VWF), platelet count, and fibrinogen were previously documented to be important predictors of critically ill patients with COVID-19 ^7-9^. A recent study revealed that specific coagulation biomarkers, such as VWF and factor VIII (FVIII) levels, are independent predictors of increased oxygen requirements in COVID[19 patients ^10^. It has also been observed that hospitalized COVID-19 patients, especially those with severe respiratory or systemic symptoms, are at increased risk for thromboembolism ^11, 12^ and aberrant bleeding manifestations ^13, 14^. Moreover, alveolar capillary microthrombi were 9 times as prevalent in patients who died from COVID-19 as those who died from H1N1 influenza ^15^. These retrospective observational studies clearly demonstrated the remarkable relevance among coagulation factor levels, thrombotic complications and COVID-19 severity. Nevertheless, it remains unclear which coagulation factor(s) can faithfully indicate the severity of COVID-19 illness or whether genetic predisposition to coagulation factor levels is causally related to severity and mortality of COVID-19 as well as the underlying biological pathways.

In the present study, by comprehensively integrating GWAS results of coagulation factors from different resources, we first utilized Mendelian Randomization (MR) analyses to investigate the causal relationships between coagulation factors and COVID-19 severity. We revealed that the antigen levels of VWF and the activities of its cleaving protease, a disintegrin and metalloproteinase with a thrombospondin type 1 motif, member 13 (ADAMTS13), causally modulate the development of severe COVID-19. Furthermore, we found polygenic risk score (PRS) of VWF is an independent clinical risk factor for predicting COVID-19 severity using UKBB COVID-19 data.

## Methods

### Data sources

#### Instrumental variables for coagulation factors

As summarized in Supplementary Table 1, we searched PubMed and GWAS Catalog ^16^ for the coagulation factor-relevant GWASs in European ethnic participants to identify genetic variants that could be used as instrumental variables for two-sample MR analysis. We selected single-nucleotide polymorphisms (SNPs) at genome-wide significant level (*P* < 5E-8), while findings showing modest significance (*P* < 5E-7) were also incorporated. The identified coagulation factors with genetic instruments include (1) Factor VIII (FVIII), Factor XI (FXI) and activated partial thromboplastin time (aPTT) that involved in intrinsic pathways; (2) Factor X (FX) and endogenous thrombin potential (ETP) that involved in common pathways; (3) Factor VII (FVII) and prothrombin time (PT) that involved in extrinsic pathways; (4) VWF and ADAMTS13 that involved in platelet adhesion; (5) D-dimer, tissue plasminogen activator (tPA), and plasminogen activator inhibitor-1 (PAI-1) concentration that involved in the dissolution of fibrin clot. Only coagulation factors with more than 3 genetic instruments were included in this study.

#### GWAS summary statistics for severe COVID-19

GWAS summary statistics for severe COVID-19 with respiratory failure were obtained from two sources, (1) the GWAS of severe COVID-19 with respiratory failure from the Severe COVID-19 GWAS Group (http://www.c19-genetics.eu), which was based on 1,610 patients and 2,205 control participants in European ethnic groups, wherein age, sex, and top 10 genetic components were adjusted ^17^; (2) the GWAS of COVID-19 with very severe respiratory confirmed from the COVID-19 Host Genetics Initiative (https://www.covid19hg.org, round 4, A2_ALL), which was based on 2,972 patients and 284,472 control participants in mixed ethnic groups (mainly from European population) ^18^.

#### UKBB COVID-19 data

The COVID-19 inspections result from the UKBB (up to 2020/10/7) was used, which included 1621 unrelated (kinship coefficient > 0.0884, corresponding to 3rd-degree relationships) UKBB participants of European ancestry (mean age 69.2 years; 53.2% men). All these participants were laboratory-confirmed COVID-19 patients. Baseline demographic and clinical characteristics of these participants are summarized in Supplementary Table 2.

The information of COVID-19 diagnosis was obtained from COVID-19 test results provided by Public Health England (PHE); death register provided by the National Health Service (NHS) Digital and NHS Central Register (NHSCR); hospital inpatient data provided by NHS Digital; and primary care data provided by TPP systems (https://www.tpp-uk.com/) and EMIS (https://www.emishealth.com/) systems. The selection criteria of UKBB participants included: ever reported as positive for SARS-CoV-2 by PHE; death from COVID-19 as the underlying cause (International Classification of Diseases, Tenth Revision (ICD-10): U071); hospitalization for COVID-19 (ICD-10: U071) from Hospital Episode Statistics; or confirmed COVID-19 infection from primary care data (Clinical Terms Version 3 (CTV3): Y20d1, EMIS: EMISNQCO303 or SNOMED CT: 1240581000000100).

Individual genotypes that primarily called by two genotyping arrays known as UKBB Axiom array and UK BiLEVE Axiom array were imputed to ∼92 million autosomal and X-chromosome variants using merged panel comprised of UK10K haplotype reference panel and 1000 Genomes Phase 3 reference panel by UKBB.

### Mendelian randomization

#### Instruments construction

The MR framework used in this study was shown in Supplementary Fig. 1. To ensure valid instruments selection according to MR analysis criteria ^19, 20^, we first harmonized instrumental SNPs for each exposure GWAS by (1) excluding SNPs associated with other potential confounders (body mass index, lipid and metabolic diseases, etc.) of exposure-outcome associations by searching PhenoScanner ^21^; (2) excluding SNPs located in the major histocompatibility complex (MHC) human leukocyte antigen (HLA) region from the list of instrumental SNPs due to potential horizontal pleiotropy with other confounder traits such as metabolic diseases and immune diseases ^22-24^; (3) excluding rare SNPs (minor allele frequency < 0.01) from the list of instrumental SNPs; (4) standardizing the effect size (β) and standard error (SE) for each SNP by the following formula ^25^:

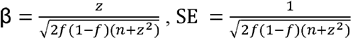

where *z* = β/SE from the original summary data, *f* is the effect allele frequency, and *n* is the total sample size.

For SNPs that were not available in the GWAS data of COVID-19 severity, we used the LDlink tool ^26^ to find the most correlated proxies (r2 > 0.8), and the summary-level statistics for proxy SNPs were used instead. Based on SNPs reported to be associated with certain exposure by the above GWAS studies, we further applied the linkage disequilibrium (LD)-based clumping implemented in PLINK ^27^ (r^2^ threshold = 0.01 and window size = 10Mb) to ensure the independence of selected SNPs. Individual-level genotype data from European population of 1000 Genomes project served as the reference panel in this study. The final SNPs used as instruments in this study are summarized in Supplementary Table 3.

#### MR statistical analysis

Several MR methodologies including Inverse variance weighted (IVW), MR-Egger regression, and weighted median (WM) methods were used to estimate the causal effects, wherein IVW was used in the main analysis ^28, 29^. We obtained Wald ratio estimates for each instrumental SNP on COVID-19 severity, and then combined Wald ratio estimates using inverse variance weighting with fixed effects. To identify potential horizontal pleiotropy, we searched PhenoScanner ^21^ to explore whether instrumental SNPs are associated with other potential cofounders of exposure-outcome associations. Moreover, we further performed several sensitivity analyses to assess the robustness of our findings by (1) we evaluated heterogeneity for causal estimates that calculated by MR-Egger regression and IVW among instrument SNPs based on Cochran’s Q statistic ^30^ and then used Mendelian Randomization Pleiotropy RESidual Sum and Outlier (MR-PRESSO) global test ^31^ to detect global pleiotropy; (2) when the global test was significant (*P* < 0.05), we removed statistically significant outliers detected by MR-PRESSO outlier test (*P* < 0.05) and repeated MR analysis; (3) to avoid multiple comparisons problem, we applied Benjamini-Hochberg method to control False discovery rate (FDR significance threshold = 0.1), and the BH-adjusted *P*-values of IVW method were used in multiple comparisons correction. IVW, MR-Egger regression, WM methods were implemented in R package TwoSampleMR v0.5.4; MR-PRESSO global test and outlier test were implemented in R package MR-PRESSO.

#### Evaluation based on UKBB COVID-19 data

Severe COVID-19 cases from the UKBB cohort were defined as laboratory- or clinical-diagnosed COVID-19 patients with at least one of the following clinical features: (1) receiving care in the intensive care unit (ICU); (2) hospitalized inpatients; (3) depending on invasive ventilation using ventilator or ventilatory supports; (4) depending on noninvasive ventilation using other enabling machines and devices. We used PRSice-2 ^32^ to construct the PRS of VWF based on the effect sizes derived from the VWF GWASs and then investigated its association with the risk of critical illness in the COVID-19 patients from UKBB cohort. PRS based on significant instrumental SNPs of VWF were calculated by the following formula ^32^:

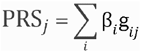

Where *j* indicates *j*^th^ individual and *i* indicates *i*^th^ variant, g is the number of risk alleles carried (*g* ∈ {0,1,2}), β is the harmonized effect size derived from the VWF GWASs.

We applied a multivariable logistic regression model to determine the association between the VWF PRS and COVID-19 severity risk with adjustments for age, sex, body mass index (BMI), systolic blood pressure (SBP), top 10 principal components of genetic variations, history of coronary artery disease (CAD), type 2 diabetes mellitus (T2DM), and chronic obstructive pulmonary disease (COPD). The significance of regression coefficients was determined by Wald statistics test under null hypothesis that the variable has no correlation with COVID-19 severity risk. To demonstrate the predictive power of PRS, we trained two logistic regression models with (1) clinical risk factors and (2) clinical risk factors + VWF PRS. Before fitting the models, we applied z-score normalization to transform raw values of variables into a same scale. We used 10-fold cross-validation in the fitting process. Specifically, we randomly divided the entire sample into 10 equal-sized sub-samples, then iteratively fitted the model using the nine folds and validated the model using the remaining one fold. Furthermore, to evaluate the fit of the model, we took the mean value of an area under the receiver operating characteristic curve (AUC) measurement among the 10 iterations.

## Results

### Causal effect of coagulation factor levels on COVID-19 severity

We systematically curated genome-wide significant SNPs associated with 12 coagulation factors from 15 GWASs, including VWF, ADAMTS13, tPA, PAI-1, D-dimer, FVII, PT, FVIII, FXI, aPTT, FX and ETP (Supplementary Table 2). The number of instrumental SNPs for each coagulation factor was summarized in Table 1. After correlated instruments removal and effect size harmonization, we performed IVW MR to test the causal effect of each coagulation factor on COVID-19 severity (Fig. 1). As summarized in Table 1, MR results based on two existing COVID-19 severity GWAS datasets showed consistent directions and comparable magnitudes of effect size. According to COVID-19 GWAS result from the Severe COVID-19 GWAS Group ^17^, among all investigated coagulation factors, we observed that VWF (*P*_IVW_ = 0.005) and ADAMTS13, also known as VWF-cleaving protease (*P*_IVW_ = 0.025), both showed significant results but displayed opposite direction of causal effect on COVID-19 severity (Table 1). Specifically, genetically determined plasma VWF antigen level was positively associated with the risk of severe COVID-19 (*P*_IVW_= 0.005, odds ratio (OR) = 1.35, 95% confidence interval (CI): 1.09-1.68) based on 17 instrumental SNPs (Supplementary Fig. 2). The significance remains after multiple testing correction (FDR = 0.06). After removing the instruments that are significantly associated with confounder traits, no additional pleiotropy was detected between VWF levels and COVID-19 severity by PRESSO global test (*P* = 0.074), Q_Egger_ (*P* = 0.777) and Q_IVM_ (*P* = 0.515). IVW MR revealed that plasma ADAMTS13 activity was inversely associated with the of severe COVID-19 (*P*_IVW_ = 0.025, OR = 0.69, 95% CI: 0.50-0.96) based on four instrumental SNPs (Supplementary Fig. 2), and no pleiotropy was detected by PRESSO global test (*P* = 0.772), Q_Egger_ (*P* = 0.433) or Q_IVM_ (*P* = 0.630). Interestingly, Given the VWF-cleaving function of ADAMTS13, this finding further supports the causal relationship between VWF levels and COVID-19 severity. However, the statistical significance disappeared after multiple testing correction (FDR = 0.15), which might be attributed to the relatively small number of valid instrumental variables.

**Figure 1.**
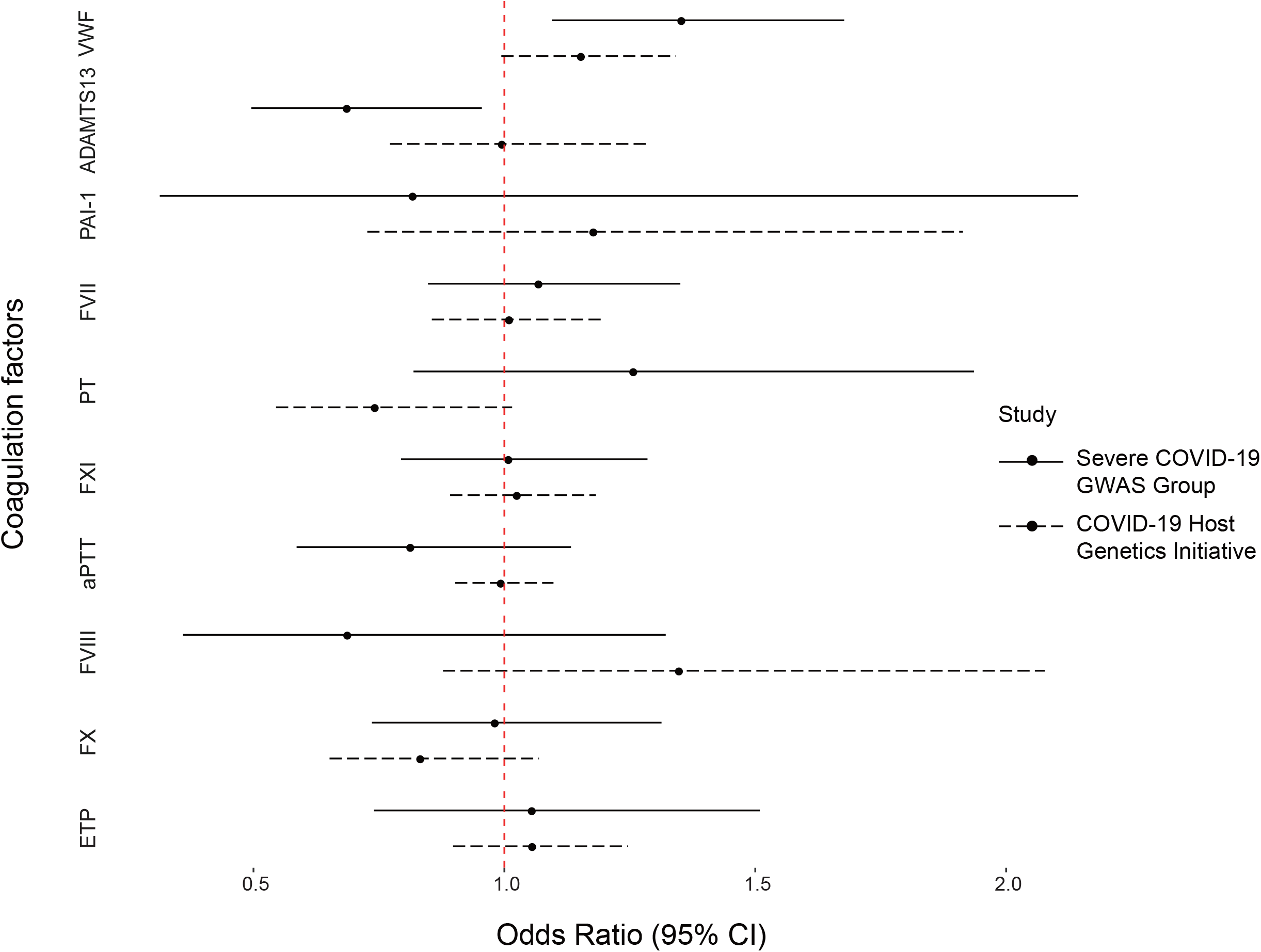
Forest plot of the associations of genetically determined coagulation factors and COVID-19 severity. Forest plot shows odds ratio (OR) and 95% confidence interval (CI) from the results of IVW MR. The solid lines indicate MR results based on COVID-19 GWAS data from the Severe COVID-19 GWAS Group ^17^ and the dashed lines indicate MR results based on COVID-19 GWAS data from COVID-19 Host Genetics Initiative ^18^. D-dimer and tPA are excluded in this plot for abnormal OR values.

**Table 1.** Summary statistics of the MR estimates of coagulation factors on COVID-19 severity.

| Exposure | nSNPs | IVW OR<br>(95% CI) | $P_{IVW}$ | Egger OR<br>(95% CI) | $P_{Egger}$ | WM OR<br>(95% CI) | $P_{WM}$ | IVW OR<br>(95% CI) | $P_{IVW}$ | Egger OR<br>(95% CI) | $P_{Egger}$ | WM OR<br>(95% CI) | $P_{WM}$ |
| --- | --- | --- | --- | --- | --- | --- | --- | --- | --- | --- | --- | --- | --- |
| The Severe COVID-19 GWAS Group <sup>17</sup> |  |  |  |  |  |  |  | The COVID-19 Host Genetics Initiative <sup>18</sup> |  |  |  |  |  |
| VWF | 17 | 1.35<br>(1.09-1.68) | **0.005** | 1.62<br>(1.24-2.13) | **0.003** | 1.47<br>(1.09-2.00) | **0.012** | 1.15<br>(0.99-1.34) | 0.060 | 1.18<br>(0.98-1.43) | 0.105 | 1.21<br>(1.01-1.46) | **0.039** |
| ADAMTS13 | 4 | 0.69<br>(0.50-0.96) | **0.025** | 0.62<br>(0.25-1.53) | 0.409 | 0.69<br>(0.48-0.99) | **0.044** | 1.00<br>(0.77-1.29) | 0.983 | 1.18<br>(0.58-2.38) | 0.697 | 1.00<br>(0.75-1.32) | 0.976 |
| D-dimer | 3 | 3.80<br>(0.52-27.90) | 0.189 | 133.44<br>(0.01-2203234.91) | 0.504 | 3.66<br>(0.28-47.28) | 0.320 | 0.61<br>(0.19-2.01) | 0.421 | 32.23<br>(0.22-4646.69) | 0.402 | 0.94<br>(0.26-3.43) | 0.925 |
| PAI-1 | 5 | 0.82<br>(0.31-2.14) | 0.685 | 0.06<br>(0.01-9.19) | 0.348 | 0.58<br>(0.17-1.92) | 0.371 | 1.18<br>(0.73-1.91) | 0.504 | 1.67<br>(0.12-22.93) | 0.726 | 1.25<br>(0.72-2.17) | 0.433 |
| tPA | 3 | 1.46<br>(0.15-14.27) | 0.743 | 1.51<br>(0.01-752.29) | 0.918 | 0.88<br>(0.14-5.75) | 0.897 | 1.42<br>(0.65-3.08) | 0.376 | 2.46<br>(0.01-618.55) | 0.803 | 1.71<br>(0.75-3.89) | 0.204 |
| FVII | 8 | 1.07<br>(0.85-1.35) | 0.569 | 0.95<br>(0.70-1.28) | 0.742 | 1.04<br>(0.81-1.34) | 0.769 | 1.01<br>(0.86-1.20) | 0.892 | 0.96<br>(0.77-1.21) | 0.767 | 0.99<br>(0.87-1.14) | 0.921 |
| PT | 6 | 1.26<br>(0.82-1.94) | 0.294 | 1.04<br>(0.51-2.11) | 0.917 | 1.13<br>(0.69-1.83) | 0.631 | 0.74<br>(0.54-1.02) | 0.062 | 0.65<br>(0.39-1.09) | 0.177 | 0.73<br>(0.50-1.06) | 0.097 |
| FXI | 3 | 1.01<br>(0.79-1.29) | 0.934 | 0.96<br>(0.59-1.58) | 0.904 | 1.01<br>(0.78-1.29) | 0.958 | 1.03<br>(0.89-1.18) | 0.711 | 0.95<br>(0.66-1.35) | 0.811 | 1.02<br>(0.89-1.18) | 0.745 |
| aPTT | 6 | 0.81<br>(0.59-1.13) | 0.223 | 1.08<br>(0.52-2.23) | 0.849 | 0.98<br>(0.80-1.20) | 0.839 | 1.00<br>(0.90-1.10) | 0.921 | 0.89<br>(0.73-1.09) | 0.315 | 0.98<br>(0.88-1.10) | 0.751 |
| FVIII | 11 | 0.69<br>(0.36-1.32) | 0.262 | 0.68<br>(0.03-17.22) | 0.819 | 0.95<br>(0.41-2.22) | 0.913 | 1.35<br>(0.88-2.08) | 0.172 | 2.71<br>(0.41-18.13) | 0.331 | 1.17<br>(0.65-2.09) | 0.601 |
| FX | 3 | 0.98<br>(0.74-1.31) | 0.908 | 1.57<br>(0.70-3.50) | 0.472 | 0.93<br>(0.67-1.30) | 0.676 | 0.83<br>(0.65-1.07) | 0.152 | 0.82<br>(0.40-1.72) | 0.697 | 0.84<br>(0.64-1.10) | 0.199 |
| ETP | 3 | 1.06<br>(0.74-1.51) | 0.761 | 1.05<br>(0.40-2.75) | 0.937 | 0.98<br>(0.73-1.31) | 0.881 | 1.06<br>(0.90-1.25) | 0.503 | 1.04<br>(0.76-1.42) | 0.851 | 1.05<br>(0.88-1.25) | 0.580 |

Values in bold indicate statistically significant results.
**Abbreviations:** **VWF**, von Willebrand factor; **ADAMTS13**, a disintegrin and metalloproteinase with a thrombospondin type 1 motif, member 13; **tPA**, tissue plasminogen activator; **PAI-1**, plasminogen activator inhibitor-1; **FVII**, Factor VII; **PT**, prothrombin time; **FVIII**, Factor VIII; **FXI**, Factor XI; **aPTT**, activated partial thromboplastin time; **FX**, Factor X; **ETP**, endogenous thrombin potential.

In addition, based on COVID-19 severity GWAS data from the COVID-19 Host Genetics Initiative round 4 ^18^, we observed that VWF is the only coagulation factor that exhibited genetic causal associations with COVID-19 severity (*P*_WM_ = 0.039, Table 1), even though there was no other MR methods to support the association except MR WM method. MR WM identified that VWF antigen level was positively associated with the risk of severe COVID-19 (*P*_WM_ = 0.039, OR = 1.21, 95% CI: 1.01-1.46). Sensitivity analyses supported the robustness of the result, where no pleiotropy was detected by PRESSO global test (*P* = 0.718), Q_Egger_ (*P* = 0.623) and Q_IVM_ (*P* = 0.680). However, no significant signal was observed from the results of ADAMTS13 MR analyses (Supplementary Fig. 2). Taken together, these results confirmed that elevated VWF is a potential causal factor for COVID-19 severity.

### The supporting evidence of VWF-ADAMTS13 as biomarkers for COVID-19 severity

A growing body of studies reported that hypercoagulation status was frequently seen in COVID-19 patients ^33, 34^. We also performed literature review to summarize existing clinical epidemiological studies regarding VWF/ADAMTS13 and COVID-19 severity. Majority of curated studies showed that the elevation of VWF antigen levels and the reduced ADAMTS13 activities are associated with COVID-19 severity (Table 2). Besides, a multi-omics analysis leveraged RNA-Seq and high-resolution mass spectrometry on 128 blood samples from COVID-19 positive and negative patients with diverse disease severities, and found VWF antigen level is significantly higher in COVID-19 patients when compared to normal controls ^35^. We also confirmed that the VWF protein level is significantly higher in ICU COVID-19 patients compared to non-ICU patients based on their released peptide quantifications (Supplementary Fig. 3). These evidences largely support that the antigen level of blood-derived VWF is an associated biomarker for COVID-19 severity.

**Table 2.** Supporting evidence for the associations between VWF/ADAMTS13 activities and COVID-19 severity.

| Study | PMID | Area | Sample size | Criteria for severity | Findings |
| --- | --- | --- | --- | --- | --- |
| Helms J <sup>69</sup> | 32367170 | French | 150 | Admitted to ICU | VWF antigen level was increased in patients with severe COVID-19 compared to healthy controls |
| Panigada M <sup>70</sup> | 32302438 | Italy | 24 | Admitted to ICU because of acute respiratory syndromes | VWF antigen level was considerably increased in ICU patients compared to healthy controls |
| Bazzan M <sup>71</sup> | 32557383 | United States | 88 | Death | Patients who died had significant lower levels of ADAMTS13 and higher levels of VWF compared to patients with non-fatal outcome |
| Krishnamachary B <sup>72</sup> | 32909001 | United States | 53 | Requires oxygen delivery by non-rebreather mask, non-invasive ventilation, or heated high flow nasal cannula at a minimum | Significantly elevated levels of VWF and decreased ADAMTS13 in large extracellular vesicles from patients with severe COVID-19 compared to healthy controls |
| Overmyer KA <sup>35</sup> | 32743614 | United States | 128 | Admitted to ICU | VWF level was increased in ICU patients versus in non-ICU patients |
| Adam EH <sup>73</sup> | 32723615 | Germany | 4 | Admitted to ICU | Elevated levels of VWF and decreased ADAMTS13 in ICU patients |
| Adrian AN <sup>74</sup> | medRxiv | Germany | 85 | The severity degree of COVID-19 was categorized according to the guidelines of the Robert Koch Institute, Germany | Elevated levels of VWF and decreased ADAMTS13/VWF ratio in severe patients |
| Goshua, G <sup>9</sup> | 32619411 | United States | 68 | Admitted to ICU | VWF antigen level was significantly elevated in ICU patients compared with non-ICU patients |

**Abbreviations:**
**ICU**, intensive care unit; **VWF**, von Willebrand factor; **ADAMTS13**, a disintegrin and metalloproteinase with a thrombospondin type 1 motif, member 13

### PRS analyses on UKBB COVID-19 cohort

According to the released COVID-19 inspection results from UKBB (up to 2020/10/7), 1,621 samples with European ethnic background were laboratory- or clinical-diagnosed as COVID-19 positive individuals. Among these positive individuals, 693 patients are classified as severe cases and remaining individuals are used as controls based on their clinical manifestations (see Methods for details). Using this independent UKBB cohort for COVID-19 severity (693 cases and 928 controls), we explored the predictive ability of PRS that derived from the VWF-associated genetic variants (17 instrumental SNPs) in the prediction of severe COVID-19 risk. Association of the VWF PRS and COVID-19 severity risk was determined using logistic regression model adjusted for age, sex, BMI, SBP, top 10 principal components of genetic variations, history of CAD, T2DM, and COPD (see Methods for details). Independent of clinical risk factors, VWF PRS was significantly associated with increased risk of severe COVID-19 (*P* = 0.047, OR per SD increase = 1.31, 95% CI: 1.00-1.71).

Previous observational studies revealed that age, sex, BMI are critical factors for predicting severe COVID-19, in addition, disease comorbidities, such as hypertension, CAD, COPD and T2DM, are also important risk factors ^2, 36-40^. To investigate whether introduction of the VWF PRS could improve the prediction performance of COVID-19 severity, we compared two logistic regression models with or without the VWF PRS by 10-fold cross validation. We found that, when the VWF PRS was added to the model, the AUC for COVID-19 severity prediction was moderately increased by 0.1%, wherein the first model using only clinical risk factors had a mean AUC of 0.699 (± 0.04) and the second model combining clinical risk factors and the VWF PRS received a mean AUC of 0.700 (± 0.04) (Fig. 2A). Since we fitted the model with z-score normalized values, the coefficients of each contributing variable can be compared directly. We observed that age is the most important risk factor for COVID-19 severity, and male sex, high BMI, and history of COPD, CAD and T2DM are also effective predictors (Fig. 2B), which is consistent with previous findings ^41-43^. Notably, VWF PRS showed larger normalized effect size than SBP (Fig. 2B), emphasizing its predictive value during the prevention and personalized treatment of COVID-19.

**Figure 2.**
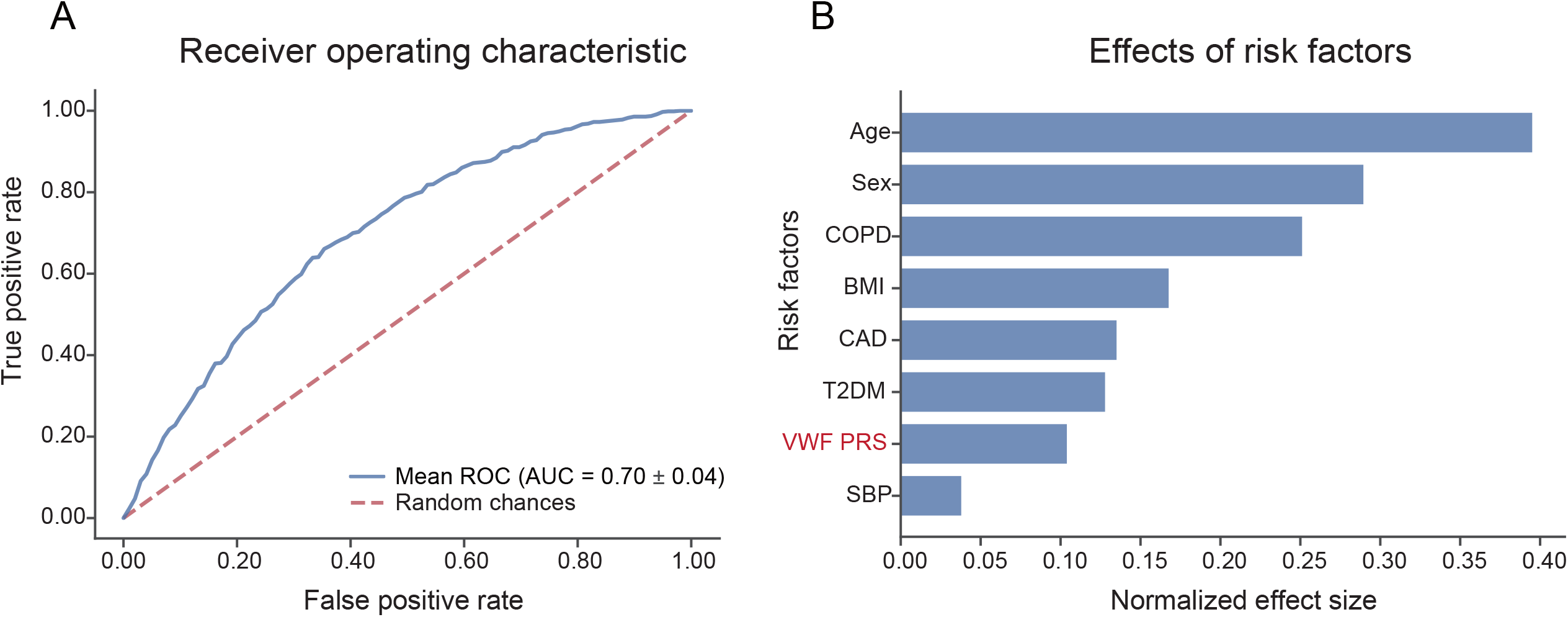
Predictive ability of VWF PRS and clinical risk factors against COVID-19 severity. (A) Receiver operating characteristic (ROC) for logistic regression using clinical risk factors and PRS derived from VWF GWAS as independent variables, area under the receiver operating characteristic curve (AUC) was the mean value for 10-fold cross validation. (B) Barplot depicts the normalized effect size of each contributing variable, values of each bar are coefficients of logistic regression after normalizing raw values to the same scale via z-score normalization. Abbreviations: **BMI**, body mass index; **CAD**, coronary artery disease; **COPD**, chronic obstructive pulmonary disease; **PRS**, polygenic risk score; **T2DM**, type 2 diabetes mellitus;

## Discussion

Emerging evidence from observational studies showed that COVID-19 patients are prone to developing thrombotic diseases ^44, 45^ which are significantly associated with COVID-19 severity, poor prognosis and mortality risk ^46, 47^. However, whether exceptional plasma levels or activities of specific coagulation factors account for a higher risk of COVID-19 severity and aberrant thrombotic manifestation is elusive. In this study, we first explored the causal relationship between multiple coagulation factors and COVID-19 severity using MR approaches. Together with the supporting evidence of recent retrospective cohort studies, our results revealed that the associations between VWF/ADAMTS13 and COVID-19 severity are essentially causal, suggesting the elevated VWF antigen and decreased ADAMTS13 activity are confident biomarkers that indicate progressive severity of COVID-19 and more aggressive critical care needed. In addition, genetic determents explain a considerable portion of the observed variance of plasma coagulation factor levels, including VWF levels ^48^. By PRS analysis on UKBB COVID-19 cohort, we uncovered that VWF PRS is an independent contributor for COVID-19 severity prediction. When combination with age, sex, BMI and several pre-existing disease statuses, the model achieved good predictive ability in distinguishing severe cases from controls.

VWF, stored in Weibel-Palade bodies and platelet α-granules for secretion upon stimulation, is a large multimeric glycoprotein which plays an important role in platelet recruitment after injury by forming a bridge between platelet surface receptors and endothelium ^49^. While, ADAMTS13 is a plasma protein cleaving VWF and decreasing its activity that anchored on the endothelial surface and in circulation ^50^. The dysfunction of VWF/ADAMTS13 dynamic equilibrium had been reported to be associated with thrombotic diseases ^51-54^ and cardiovascular diseases ^55^. COVID-19 patients are prone to developing thrombotic diseases ^44, 45^, reciprocally, the development of thrombotic diseases could account for poor prognosis and mortality of COVID-19 ^46, 47^. We suspected that SARS-COV-2 invades human lungs and causes injury or inflammation in the blood vessels, which then promoting a pro-coagulative state. As an important consideration for COVID-19, elevated VWF level and insufficient ADAMTS13 activity confer a higher risk of forming blood clots and ultimately develop venous thromboembolism. Thrombus can block normal blood flow and decrease oxygen supplement to alveoli, and this may explain partially why COVID-19 patients are at risk of respiratory failure. According to our MR causal inference and PRS analysis in the UKBB COVID-19 cohort, we could advise individuals carry certain VWF/ADAMTS13 alleles to closely monitor their coagulation parameters and take supportive care for coagulopathy prevention after SARS-CoV-2 infection. Also, existing drugs targeting VWF or its molecular interactions could be considered to control COVID-19 severity and associated thrombotic complication for specific cohort. Several other coagulation factors such as D-dimer and PT were previously documented to be associated with COVID-19 severity in observational studies ^7, 8^, however, we found no causal relationships through MR analyses. This phenomenon could be attributed to potential confounding factors in observational studies, where associations between coagulation factors and COVID-19 severity was not due to direct causality, but rather because both coagulation factors and COVID-19 severity are likely caused by other unrevealed confounders.

*ABO* gene loci obtained the most prominent GWAS signal in plasma VWF levels ^48^ and strong associations with COVID-19 ^17^, implying ABO blood group may confound the establishment of causality between VWF/ADAMTS13 and COVID-19 severity. Recent epidemiological studies have investigated and observed tight association of ABO blood group with the COVID-19 susceptibility, severity and mortality ^56-59^. It was also reported that the *ABO* genotypes and ABO blood group are associated with ACE activity in hypertensive patients ^60^, and increased plasma ABO protein is causally associated with the risk of severe COVID-19 ^61^. Among the associated variants of VWF levels within the *ABO* locus, SNPs rs10901252 and rs687621 can perfectly discriminate B and O blood groups from A ^62^. Individuals with blood group O have lower VWF plasma concentrations compared with individuals with blood group non-O ^63^. The presence of blood group A and B antigens on VWF molecules may have clinically significant effects on VWF proteolysis and clearance ^64^. In our analysis, we excluded SNP rs687621 due to potential horizontal pleiotropy with other traits (such as Interleukin 6 levels and coronary artery disease) and rs10901252 for high LD with other instrumental SNPs (such as rs8176743). We found that the causality between VWF levels and COVID-19 severity was established when taking away the genetic effect of these SNPs. However, whether the causal mechanisms of VWF levels on COVID-19 severity could be independent of ABO blood group still needs further investigation.

Our study is also subject to some data limitations. First, only genome-wide significant variants were available from existing GWAS results of VWF/ADAMTS13 and most of investigated coagulator factors, which makes it impossible to perform bi-directional MR analysis, and such limited number of instrumental SNPs for particular coagulation factors could affect the accuracy of both MR and PRS estimations. Similar to recent PRS studies on CAD ^65^ and ischemic stroke ^66^, we observed that incorporation of genetic component significantly contributes the prediction model but only slightly improves the overall performance. The significant association between VWF PRS and COVID-19 severity indicated that the information captured by VWF PRS is not fully explained by other risk factors. But the current PRS study on COVID-19 cohort is likely underpowered due to insufficient sample size, the borderline significance required a larger study to ascertain the accuracy. Second, genetic risk loci for coagulation factors may vary among different populations ^67, 68^, but our study mainly focuses people from European descent. Whether there are population-specific causal mechanisms needs further exploration. Also, we didn’t investigate gender- or age-specific effects because of the lack of gender- or age-stratified GWAS data. Last, the sample size of severe COVID-19 GWAS is still insufficient in current stage, and further research is warranted when abundant and non-European ethnic GWAS data is available in the future. We also expect that prospective controlled trial could be applied to ascertain the causal role of VWF/ADAMTS13 and investigate potential treatments for certain infected populations.

## Data Availability

1. Explicit statement that approval was received to use the data in the present work or that the access to the data was granted after registration.
#Response: GWAS summary data from the Severe COVID-19 GWAS Group and the COVID-19 Host Genetics Initiative are publicly available. The UKBB COVID-19 information including genotypes, individual clinical information are granted by UKBB, the approved project is "Identification and validation of causal variants effect on cardiovascular disease through immune-mediated inflammation".
2. Additional details (e.g. approval code / registration ID#), if applicable
#Response: The UKBB COVID-19 project granted ID: 44482.
3. Statement about the ethics oversight body that gave ethical approval for the collection of the original data.
#Response: All ethical approval for the collection of the original data is declared by UKBB and we comply with the UKBB data usage restriction.

## Acknowledgements

Part of the data used in the analyses described in this manuscript were obtained from the UKBB and public GWAS resources. We appreciate all resource and tool providers.

## Funding

This work was supported by grants from the National Natural Science Foundation of China 32070675, 31871327 (M.J.L.), Natural Science Foundation of Tianjin 19JCJQJC63600 (M.J.L.).

## Competing interests

The authors declare that they have no competing interests.

## Notes

### Competing Interest Statement

The authors have declared no competing interest.

### Author Declarations

GWAS summary data from the Severe COVID-19 GWAS Group and the COVID-19 Host Genetics Initiative are publicly available. The UKBB COVID-19 information including genotypes, individual clinical information are granted by UKBB, the approved project is "Identification and validation of causal variants effect on cardiovascular disease through immune-mediated inflammation", project ID: 44482

### Summary of Updates

author update author affiliations updated manuscript main text revised figure 1 and figure 2 revised

